# The role of tuberculosis symptoms in transmission risk to cell contacts in prisons

**DOI:** 10.1101/2025.05.27.25328413

**Authors:** Wanessa da Silva Peres Bezerra, Esther Jung, Mariana G. Croda, Roberto Dias de Oliveira, Andrea da Silva Santos, Daniel Henrique Tsuha, Paulo Cesar Pereira Dos Santos, Eunice Atsuko Totumi Cunha, Julio Croda, Jason R. Andrews

## Abstract

**Background:** Understanding determinants of *Mycobacterium tuberculosis* transmission is critical to devising effective strategies to reduce its burden. Whether and to what extent symptoms influence transmission remains poorly understood.

**Methods:** Between 2020 and 2022, we systematically screened PDL from three prisons in Brazil for TB by symptom assessment and sputum testing with Xpert MTB/RIF Ultra. We performed QuantiFERON-TB Gold Plus (QFT) testing among cell contacts of individuals with TB and in cells with no TB case identified. We evaluated the relationship between TB exposure (symptomatic, asymptomatic, none) and QFT positivity using Bayesian generalized linear mixed models.

**Results:** We screened 7641 PDL for TB and identified 290 cases, yielding a prevalence of 3.8% (290/7641). After applying exclusion criteria, 686 participants were included for QFT analysis: 132 contacts of 42 individuals with symptomatic TB, 224 individuals exposed to 52 individuals with asymptomatic TB; 330 individuals with no recent cell exposure. Odds of QFT positivity were higher in symptomatic (adjusted odds ratio [aOR] 2.50, 95% CrI 1.51–4.16) and asymptomatic (aOR 1.61, 95% CrI 1.06–2.45) exposure groups compared to those unexposed. QFT positivity in contacts of symptomatic and asymptomatic TB did not differ (aOR 1.56, 95% CrI 0.92-2.63).

**Conclusions:** Over half of individuals with TB lacked symptoms, and their contacts had increased risk of QFT positivity—comparable to contacts of symptomatic TB— compared with individuals without recent cell exposure. These findings underscore the importance of systematic screening for TB, irrespective of symptoms, to accelerate diagnosis and prevent transmission in high-burden settings.

**Article Summary:** An active case finding program in Brazilian prisons identified high burden of asymptomatic, microbiologically confirmed TB, and contacts of tuberculosis cases without symptoms had similarly elevated risk of *Mtb* infection as contacts of symptomatic cases.

## INTRODUCTION

TB remains the leading cause of death from a single infectious disease, with over 10 million new cases and 1.25 million deaths estimated to have occurred in 2023^1^. One of the major obstacles to eliminating TB as a public health threat is the ongoing transmission of *Mycobacterium tuberculosis* (*Mtb*), resulting in millions of new infections every year. Despite this, determinants of TB infectiousness remain poorly understood.

Recent studies have shown that approximately half of individuals living with undiagnosed TB lack symptoms^2^, yet the contribution of this group to transmission remains uncertain. An analysis of household contacts of individuals with TB found that individuals without symptoms were as infectious as those with symptoms^3^. Combining these findings with data on the prevalence of asymptomatic and symptomatic TB from prevalence surveys, it was estimated that 68% of all *Mtb* transmission arises from individuals who lack symptoms^4^. However, as prior studies have estimated that the majority of *Mtb* infections are acquired outside the household^5^, it is possible that determinants of infectiousness measured among putative index cases in households might be biased towards the null.

Globally, prisons have among the highest incidence rates of TB and very high rates of *Mtb* transmission. A recent meta-analysis estimated that TB incidence in South American prisons is 26 times higher than that of the general population^6^. Prior studies in prisons in central western Brazil, where community rates of TB transmission are modest, found that less than 10% of individuals entering prisons test positive for TB infection^7^; however, 25-50% of individuals with a negative tuberculin skin test (TST) or interferon gamma release assay convert to a positive test each year^8,9^. Further, exposure to a case within a cell is associated with the risk of TST conversion, with a risk of conversion among contacts of smear positive cases three times greater than that of contacts of smear negative cases.

We sought to understand the role of asymptomatic TB in transmission by performing active case finding in three high TB burden prisons and comparing QuantiFERON-TB Gold Plus (QFT) positivity rates among individuals residing in the same cell as individuals with symptomatic or asymptomatic TB or with no defined exposure.

## METHODS

### Overview

We performed systematic screening for TB in three high TB burden prisons in Mato Grosso do Sul, Brazil, each housing over 1,000 incarcerated persons. Mato Grosso do Sul, located in Central-West Brazil and bordering Paraguay and Bolivia, has the highest incarceration rate in the country, driven primarily by drug-trafficking offenses.

Between August 2020 and November 2022, study nurses recruited individuals across the three prisons and obtained informed consent. Screening was offered to all residents of the three prisons, and more than 95% provided informed consent and agreed to participate. After obtaining informed consent, a standardized questionnaire was administered to all participants to elicit demographic and clinical information, including TB symptoms. TB-related symptoms were defined as cough of any duration, sputum, fever, appetite loss, night sweats, chest pain, trouble breathing, and weight loss of any duration. All consenting participants, regardless of symptoms, had a spot sputum sample collected for testing using the Xpert^®^ MTB/RIF Ultra assay.

### Study Design and Population

A TB case was defined by a positive sputum test by Xpert® MTB/RIF Ultra. Trace results were repeated with a new sample. Those that persisted as traces were considered negative for TB. Cases were further classified as symptomatic or asymptomatic TB based on their responses to the symptom questions during the mass screening. Symptomatic TB was diagnosed in individuals presenting with one or more of the specified TB symptoms, irrespective of their duration, while asymptomatic TB referred to cases without any of these TB symptoms.

To recruit contact participants, we reviewed the list of individuals sharing a cell with each individual with TB to identify eligible individuals. Individuals that had negative sputum Xpert, who had been residing in a cell with an individual with TB for at least two weeks before the index case’s diagnosis, who had no prior TB diagnosis, and who had not previously been incarcerated were eligible for inclusion. Individuals residing in cells with two or more TB cases were excluded. Additionally, an unexposed group was recruited following the same criteria, except that they were selected based on residing in cells in which no one was found to have TB.

We originally aimed to recruit 1356 contact participants to have 90% power to detect an odds ratio of 0.5 or lower comparing the exposure groups (symptomatic versus asymptomatic), assuming a large design effect (2.5) from sampling multiple individuals per cell with correlated outcomes. We recruited and screened 1,645 participants; however, due to the high prevalence of incarceration history among screened individuals, the final analytic sample consisted of 686 eligible participants (Figure 1): 132 PDL exposed to a symptomatic TB index case in the cell, 224 were exposed to an asymptomatic TB index case, and 330 unexposed to a TB case in the cell. Recruitment was concluded once it became clear that further enrollment of eligible individuals was not feasible within the constraints of the study period and setting. As the design effect was substantially lower than anticipated, this sample size provided 80% power to detect a OR <0.5.

**Figure 1.**
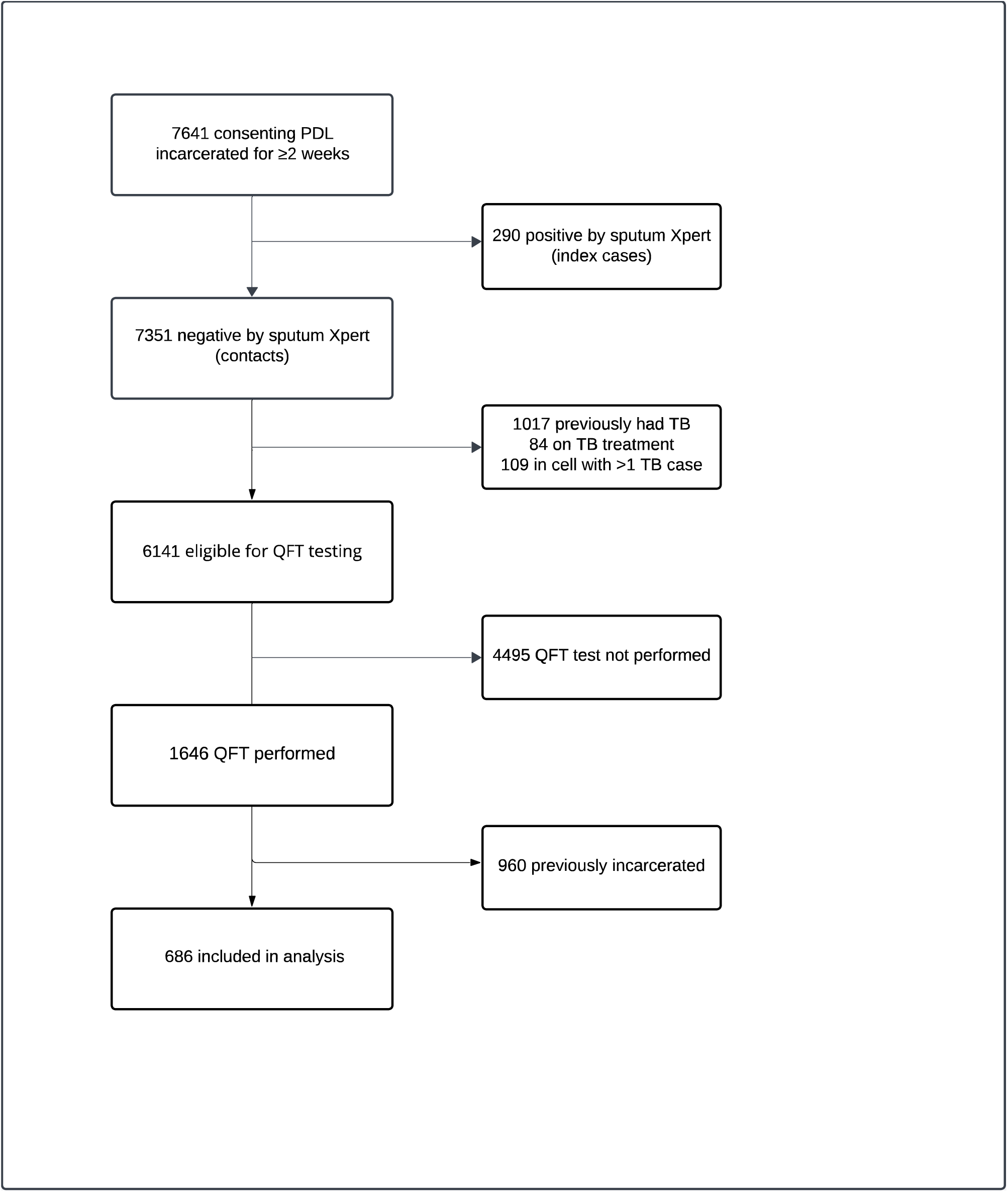
Overview of the study population

Peripheral venous blood sample was collected from all eligible participants for QFT testing and performed according to the manufacturer recommended instructions (Qiagen, Venlo, The Netherlands). QFT positivity was evaluated using the manufacturer-recommended threshold of TB antigen minus nil ≥0.35 IU/mL in either tube. Sensitivity analyses were conducted using ≥0.70 IU/mL and ≥4.00 IU/mL thresholds, following previous publications^10,11^.

### Statistical Analysis

Generalized linear mixed models (GLMMs) were used to compare QFT positivity across the defined exposure categories. Fixed effects included the TB exposure categories, Xpert semiquantitative results (negative, low/very low, medium/high), age, prison, number of cellmates, and months incarcerated. Random effects accounted for cell-level clustering.

Bayesian hierarchical models were fitted using the ‘stan_glmer’ function in the rstanarm package (version 2.31.1)^12^ with default prior specifications. Monte Carlo Markov Chain (MCMC) sampling was performed with four chains, each comprising 2,000 iterations and a warm-up of 1,000 iterations, to estimate adjusted odds ratios (aORs) with 95% credible intervals (CrIs). All analyses were conducted in R (version 4.4.1) with a random seed of 123 for reproducibility. Analytic code is available at https://github.com/Andrews-Lab-Stanford/AsymptomaticTB-BrazilPrisons/releases/tag/v1.0.

### Ethical Considerations

This study was approved by the National Commission for Ethics in Research (CONEP) (3.483.377, CAAE 37237814.4.0000.5160), the Federal University of Mato Grosso do Sul Research Ethics Committee (CEP), and Stanford University. All participants provided written informed consent prior to enrollment.

## RESULTS

### Systematic tuberculosis screening and index case identification

From August 2020 to November 2022, we screened 7641 individuals for TB and identified 290 individuals with TB confirmed by a positive sputum Xpert result. Among these, 104 (35.9%) reported any TB symptoms. Individuals with symptomatic and asymptomatic TB had similar ages, education levels, substance use patterns (smoking, alcohol, and illicit drug use), and exposure to TB-related risk factors, including knowing someone with TB and a history of incarceration (Table 1).

**Table 1.**
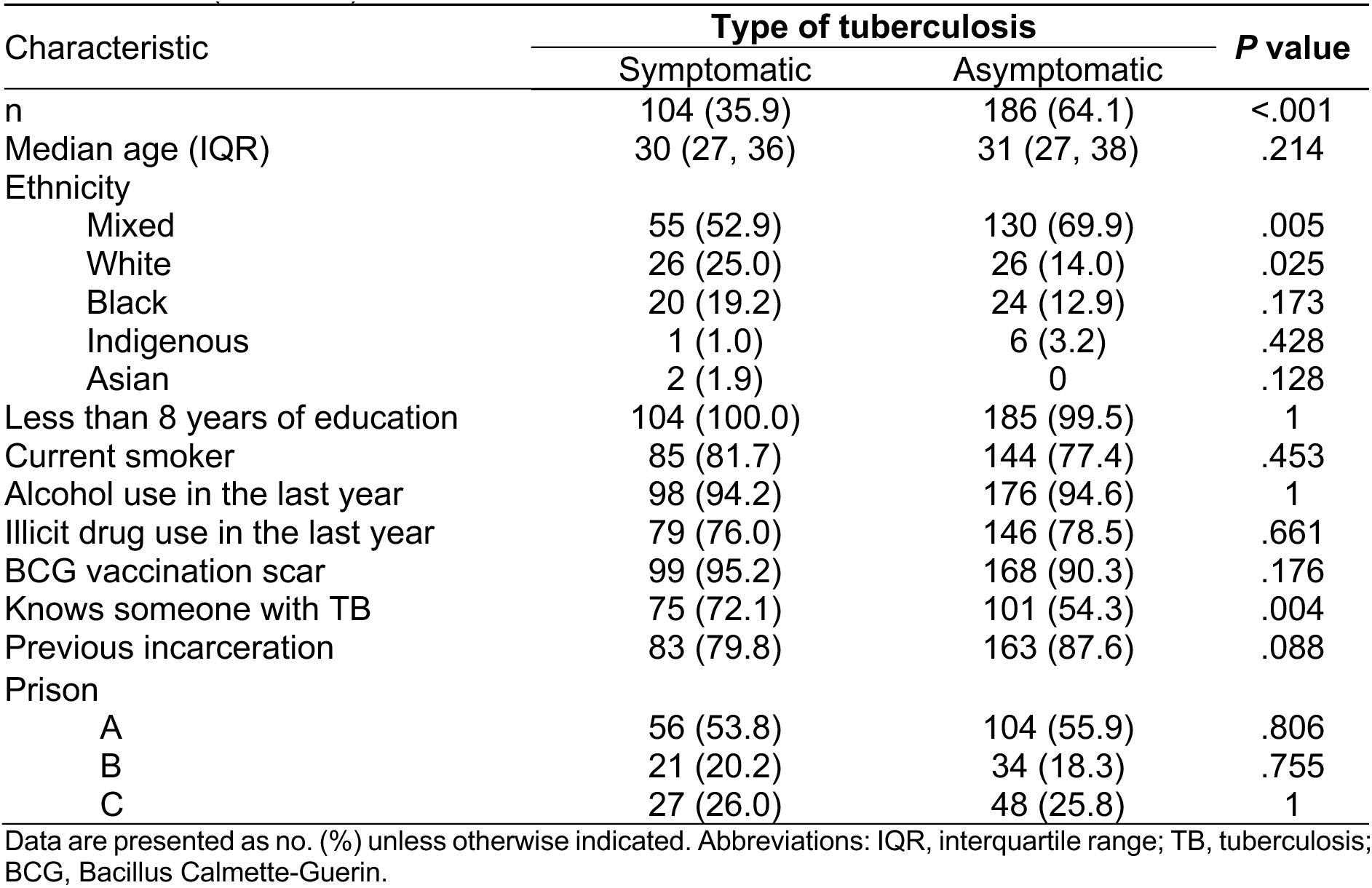
Demographic and clinical characteristics of index cases by type of tuberculosis (N = 290)

**Table 2.** Demographic and clinical characteristics of participants by type of exposure to TB in the cell (N = 686)

| Characteristic | Tuberculosis exposure classification |  |  | P value |  |  |
| --- | --- | --- | --- | --- | --- | --- |
|  | Symptomatic | Asymptomatic | No exposure | Symptomatic/<br>Asymptomatic | Symptomatic/<br>No exposure | Asymptomatic/<br>No exposure |
| n | 132 | 224 | 330 |  |  |  |
| Median age (IQR) | 31 (24, 39) | 35 (27, 60) | 30 (25, 40) | <.001 | .987 | <.001 |
| Ethnicity |  |  |  |  |  |  |
| Mixed | 79 (59.8) | 121 (54.0) | 187 (56.7) | .320 | .603 | .543 |
| White | 38 (28.8) | 66 (29.5) | 116 (35.2) | .905 | .229 | .168 |
| Black | 13 (9.8) | 27 (12.1) | 20 (6.1) | .604 | .164 | .019 |
| Indigenous | 2 (1.5) | 7 (3.1) | 5 (1.5) | .494 | 1 | .240 |
| Asian | 0 (0.0) | 3 (1.3) | 2 (0.6) | .298 | 1 | .398 |
| Less than 8 years of education | 130 (98.5) | 213 (95.1) | 311 (94.2) | .143 | .050 | .707 |
| Current smoker | 58 (43.9) | 118 (52.7) | 172 (52.1) | .125 | .123 | .931 |
| Alcohol use in the last year | 8 (6.1) | 14 (6.2) | 15 (4.5) | 1 | .485 | .438 |
| Illicit drug use in the last year | 59 (44.7) | 66 (29.5) | 138 (41.8) | .004 | .603 | .003 |
| BCG vaccination scar | 120 (90.9) | 178 (79.5) | 310 (93.9) | .005 | .310 | <.001 |
| Knows someone with TB | 77 (58.3) | 88 (39.3) | 66 (20.0) | .001 | <.001 | <.001 |
| Months incarcerated (IQR) | 152 (75, 397) | 187 (83.5) | 200 (76, 342) | .003 | .016 | .546 |
| Number of individuals in cell |  |  |  |  |  |  |
| ≤ 10 | 12 (9.1) | 33 (14.7) | 68 (20.6) | .139 | .003 | .092 |
| 11-20 | 56 (42.4) | 46 (20.5) | 203 (61.5) | <.001 | <.001 | <.001 |
| 21-40 | 23 (17.4) | 83 (37.1) | 43 (13.0) | <.001 | .240 | <.001 |
| 41-60 | 41 (31.1) | 62 (27.7) | 16 (4.8) | .546 | <.001 | <.001 |
| Prison |  |  |  |  |  |  |
| A | 43 (32.6) | 40 (17.9) | 51 (15.5) | .002 | <.001 | .484 |
| B | 36 (27.3) | 57 (25.4) | 201 (60.9) | .710 | <.001 | <.001 |
| C | 53 (40.2) | 127 (56.7) | 78 (23.6) | .003 | .001 | .242 |

Data are presented as no. (%) unless otherwise indicated. Abbreviations: IQR, interquartile range; TB, tuberculosis; BCG, Bacillus Calmette-Guerin.

### Study population by exposure status

Among the 7351 Xpert-negative PDL, 1017 were excluded due to a history of previous TB and 84 were excluded because they were undergoing TB treatment. A total of 6141 Xpert-negative inmates, who met the inclusion criteria of residing in the same cell for at least two weeks, residing in a cell without more than one TB case, with no history of TB, and not receiving treatment for TB, met initial eligibility criteria for the exposure study. Of these individuals, 1645 were recruited for the study, and 960 were subsequently excluded due a self-reported history of previous incarceration, resulting in a final study population of 686 participants who underwent QFT testing: 132 exposed to symptomatic TB, 224 exposed to asymptomatic TB, and 330 without recent known exposure to TB.

Demographic and clinical characteristics were similarly distributed and balanced across the three prisons, with a few significant differences. Participants exposed to a person with asymptomatic TB had a higher median age (35 years [IQR: 27–60]) compared to the symptomatic exposure (31 years [IQR: 24–39]; p < .001) and no-exposure group (30 [25–40]; p < .001) (Table 1). Educational attainment was low across all groups, with over 94% having less than eight years of schooling. Approximately half of the participants reported being smokers, with no significant differences between exposure groups. BCG vaccination scars were significantly less common in the asymptomatic exposure group (79.5%) than in the symptomatic exposure (90.9%; p = .005) and no-exposure groups (93.9%; p < .001).

Several carceral characteristics differed between the exposure groups. The median duration of incarceration was longer in the asymptomatic exposure (187 months, p = .003) and no-exposure groups (200 months, p = .016) compared to the symptomatic exposure group (152 months). The number of individuals in each cell differed significantly: 49% of participants with a symptomatic exposure and 65% of those with an asymptomatic exposure resided in cells with more than 20 PDL, compared with 18% of participants with no TB exposure (p < .001).

### Predictors of QuantiFERON positivity

At the >0.35 IU/mL threshold, 74% of participants exposed to symptomatic cases, 65% of participants exposed to asymptomatic cases, and 46% of unexposed participants were QFT-positive. The association between QFT positivity and incarceration-related factors was assessed across the three prisons (Figure 2). QFT positivity rates increased with prolonged incarceration across all prisons (Figure 2A). Higher cell density, defined as the number of individuals per square meter in each cell, increased with QFT positivity across all prisons (Figure 2B), emphasizing the role of overcrowding in facilitating TB transmission.

To evaluate the relationship between QFT positivity and TB exposure, mixed effects logistic regression analysis was performed with fixed effects for TB exposure status (unexposed, asymptomatic, symptomatic), age, prison, number of cellmates, and duration of incarceration and random effects for the cell. Odds of QFT positivity were significantly higher among individuals with exposure to symptomatic cases (aOR 2.50; 95% CrI 1.51–4.16; p < .001) or asymptomatic cases (aOR 1.61; 95% CrI 1.06–2.45; p = .026) relative to those without TB exposure (Table 3). Odds of QFT positivity were not significantly different between those exposed to symptomatic cases (aOR 1.56; 95% CrI 0.92-2.63; p = .099) and asymptomatic cases (Table 3). Adjusting for exposure type, duration of incarceration remained positively associated with QFT positivity (aOR: 1.01 per month; 95% CrI: 1.01–1.02; p < .001). Cell density was also positively associated with QFT positivity (aOR: 1.81; 95% CrI: 1.16–2.84; p = .009) (Table 3).

**Table 3.**
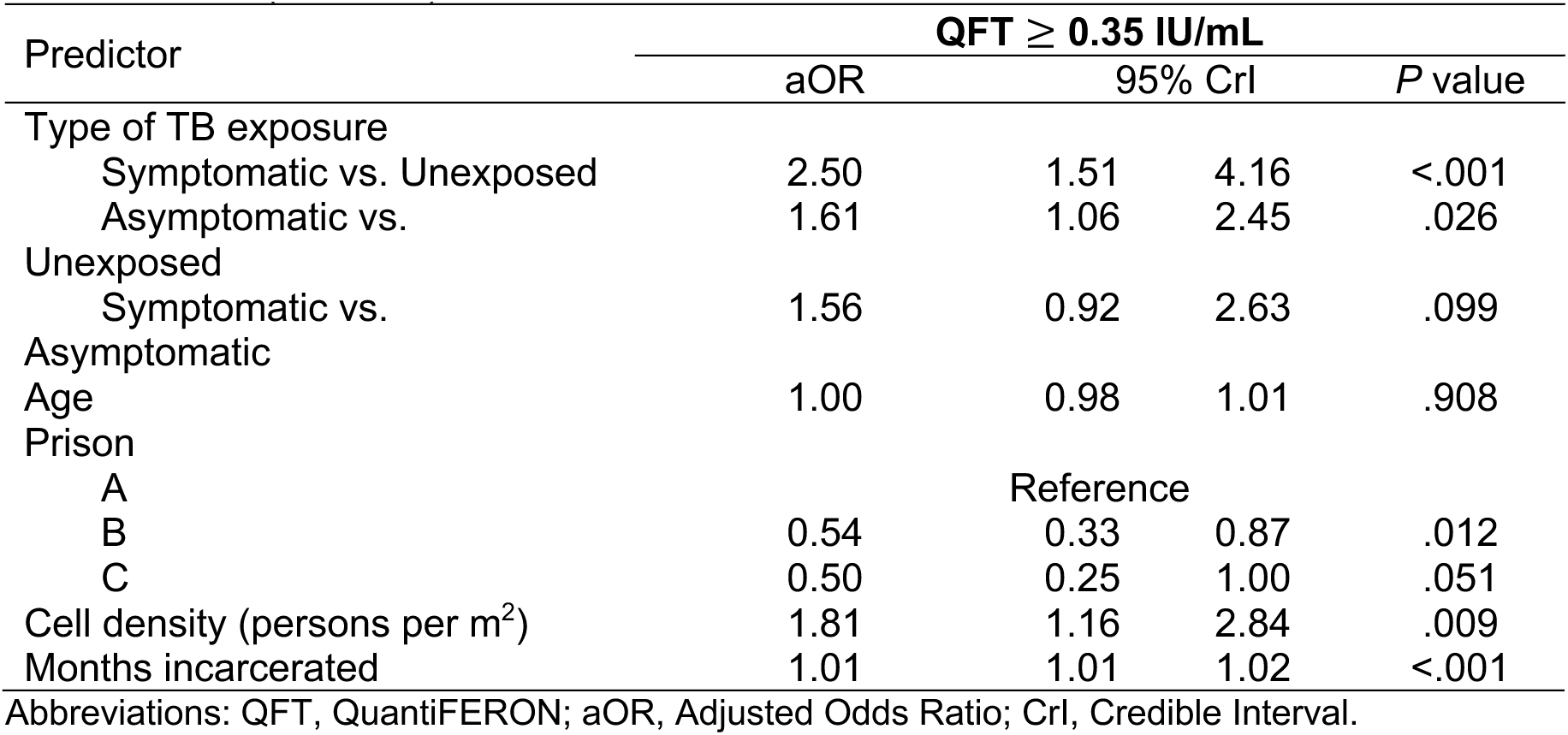
Mixed effects logistic regression analysis of QFT positivity (≥0.35 IU/mL) evaluating effect of tuberculosis exposure type among individuals with no previous incarceration (N = 686)

Participants exposed to an individual with very low or low Xpert semi-quantitative sputum bacillary load had higher odds of QFT positivity (>0.35 IU/mL) compared to unexposed participants (aOR 1.61; 95% CrI 0.99–2.63; p = .056). Those exposed to individuals with medium or high Xpert semi-quantitative sputum bacillary load had higher odds of QFT positivity (aOR: 2.34; 95% CI: 1.42–3.84; p < .001), but the differences in QFT positivity between those exposed to medium/high compared with low/very low bacillary load were not statistically significant (aOR: 1.44; 95% CI: 0.82–2.53; p = .204) (Table 4).

**Table 4.**
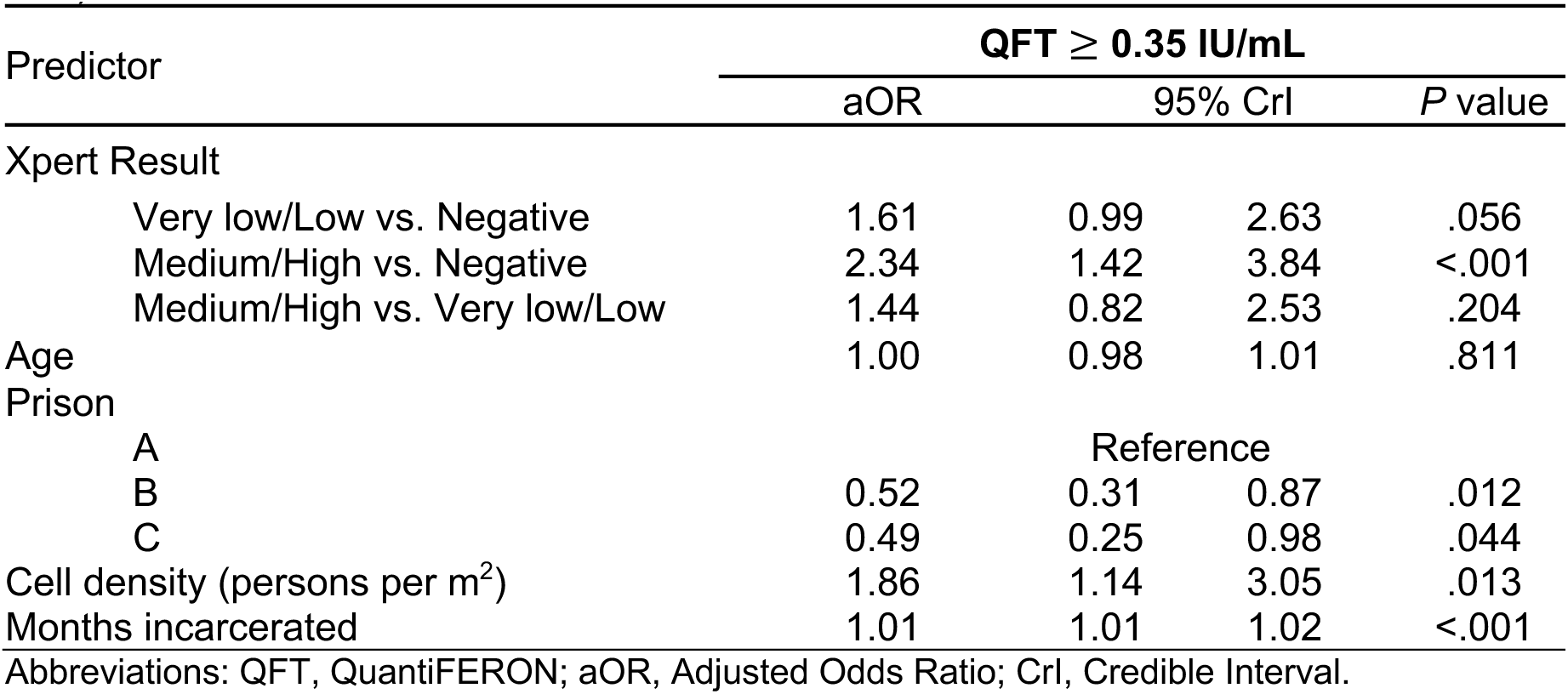
Mixed effects logistic regression analysis of QFT positivity (≥0.35 IU/mL) evaluating effect of Xpert result among individuals with no previous incarceration (N = 686)

### Sensitivity analyses

Sensitivity analyses at higher QFT positivity thresholds (>0.70 IU/mL and >4.0 IU/mL) yielded similar findings (Supplementary Tables 1 and 2). Likelihood of high QFT positivity (>4.0 IU/mL) was higher among symptomatic contacts (aOR: 3.35, 95% CrI 1.55–7.24, p = .002) and asymptomatic contacts (aOR: 3.00; 95% CI: 1.42– 6.36; p = .004). Duration of incarceration was significantly positively associated with QFT positivity at all thresholds. Medium or high Xpert results were associated with a substantial increase in the odds of QFT positivity (>4.0 IU/mL) (aOR: 3.55; 95% CI: 1.67–7.54; p = .001) compared to negative results. Similarly, very low or low Xpert results were also significantly associated with increased odds of QFT positivity (>4.0 IU/mL) (aOR: 2.74; 95% CI: 1.26–5.95; p = .011) compared to negative results.

## DISCUSSION

Around half of all individuals with prevalent TB lack symptoms^2^, and it is critical to understand their contribution to *Mtb* transmission in order to develop optimal case detection strategies. Through systematic screening for TB and *Mtb* infection in prisons, we found that individuals residing in cells with persons with symptomatic or asymptomatic TB had increased risk of *Mtb* infection compared to those residing in cells without an identified case. We did not find differences in risk between the groups exposed to TB according to symptom status. These findings were robust to the definition of QuantiFERON positivity, with stronger associations at more stringent thresholds. These findings underscore the importance of systematic screening, including among individuals who lack TB symptoms, in high burden settings such as prisons.

Nearly two-thirds (64%) of individuals identified to have microbiologically-confirmed TB in this study were asymptomatic at screening. This high proportion of asymptomatic TB identified through active case finding is consistent with prior studies performed in prisons^7,13^ and in community settings^4^. Systematic screening with more sensitivity modalities such as chest radiography or sputum pooling can improve case detection while reducing costs compared with individual testing of all individuals with molecular diagnostics^14–16^. Beyond diagnostics, addressing structural determinants in prisons, including overcrowding and inadequate ventilation, remains critical^17,18^. Significant differences in QFT positivity among the three prison suggest that structural and environmental factors influence *Mtb* transmission, and that these can be modifiable. Further studies are needed to identify scalable structural or environmental interventions to reduce *Mtb* transmission in prisons and other congregate settings.

There have been fewer data concerning the infectiousness of asymptomatic TB. A recent analysis of population surveys from three countries in Asia in which household tuberculin skin test or IGRA data were available found no difference in odds of infection in contacts of symptomatic versus asymptomatic cases (aOR 1.2, 95% CI 0.6-2.3)^3^. Our findings, from a very different population and environment, are nevertheless consistent with this result. We found that contacts of both asymptomatic and symptomatic TB had elevated infection risk, but the risk in contacts of symptomatic TB was not statistically greater (aOR 1.56; 95% CrI 0.92-2.63) than that in contacts of asymptomatic TB. While the Emery study further found that contacts of individuals with smear positive TB had greater odds of *Mtb* infection^3^, we did not have smear data available as it is no longer used in this setting. We evaluated the influence of semi-quantitative Xpert result, a measure of bacillary load, in the index case on risk of *Mtb* infection in contacts and found a non-significant trend, with 1.44-fold greater odds in those exposed to a medium or high bacillary load compared with a low or very low load.

One of the difficulties in interpreting QuantiFERON results is that positivity is very sensitive to the threshold for the antigen minus nil response, and there is a lack of bimodality to guide this threshold selection. Several studies have shown that higher values provide greater positive predictive value for TB risk and lower probability of reversion^10,19–21^. We evaluated two alternative, higher thresholds to define positivity (>0.70 IU/mL and >4.00 IU/mL) and found that the associations between TB exposure, symptoms and odds of QuantiFERON positivity were robust to the threshold utilized.

A major limitation to cross-sectional studies assessing the relationship between TB exposure and *Mtb* infection is that it is unknown when infections occurred. Exposures that are not due to the index case in these studies (i.e., infections acquired outside the household or prison cell) bias the effect estimates for index characteristics towards the null. In household studies in high burden communities, it is estimated that 80% or more of infections are acquired outside of the household^22^. In prisons, the proportion of infections acquired in the cell is not known. Our studies in Brazil have found that less than 10% of individuals have positive tuberculin skin tests at the time of first entry into the prison^7^. Individuals are in their cells for 20-21 hours per day, with only 3-4 hours of recreational time, which occurs in open air courtyards. Together, these suggest that in-cell exposures are important for *Mtb* infection, and evidence from genomic clustering of TB isolates supports this^23^. However, individuals are transferred between cells frequently. We performed sensitivity analyses on total duration of incarceration and cell crowding and the results were robust to these specifications (Figure 2). Intensive cohort studies, in which QuantiFERON-negative individuals are following longitudinally for risk of infection, could mitigate some of these difficulties in relating exposures to infections, but will be more resource-intensive to perform.

Another difficulty with comparing *Mtb* transmission risk according to symptom status is that symptoms are dynamic, but measurements are typically only obtained at the time of diagnosis. It is presumed that individuals with symptomatic TB at some point had asymptomatic disease^24–27^, but the duration that individuals spend in that asymptomatic state likely varies considerably. Further, symptoms can regress, such that individuals with asymptomatic TB at time of diagnosis might have had symptoms—whether due to TB or another cause—at some point during their disease course. Longitudinal monitoring technologies might be one solution to overcome this, but given that such monitoring would have been initiated before an individual develops disease, the number of participants needed to include in such studies would be prohibitive with our present technologies.

Another limitation of this study is we were unable to assess the impact of HIV on transmission. HIV test results were available for only around 50% of contacts. Among the 3,811 individuals who were tested, the HIV positivity rate was 0.9% (35/3,811), which is consistent with the relatively low prevalence reported in previous studies conducted in prisons in the state of Mato Grosso do Sul^8,28,29^.

In summary, we found that individuals exposed to asymptomatic TB—which comprised the majority of prevalent TB in this study—have QFT positivity comparable to those exposed to symptomatic cases and higher than those without recent *Mtb* exposure. These findings affirm an important role for asymptomatic TB in transmission, confirming the need to use symptom-agnostic screening modalities such as chest radiography or sputum pooling. Given the extremely high burden of TB in prisons globally, frequent systematic screening to identify all individuals with TB will be critical for averting transmission and reducing incidence.

## Data Availability

All data produced in the present study are available upon reasonable request to the authors

## Acknowledgements

JRA and JC conceived of the study. JRA, JC, MGC, RDD and EATC developed the study plans. WdSPB enrolled participants and collected data. PCPdS and EATC performed laboratory testing and DHT performed data management. WdSPB, EJ and JRA performed data analysis and wrote the manuscript. All authors contributed to revising the manuscript.

## Funding

This work was supported by Brazil’s Ministry of Health; Brazil’s National Council for Scientific and Technological Development [CNPq; 404182/2019-4 to JC] and the U.S. National Institutes of Health [R01 AI130058, R01 AI149620, K24AI182647 to JRA].

## Notes

### Competing Interest Statement

The authors have declared no competing interest.

### Clinical Protocols

https://github.com/Andrews-Lab-Stanford/AsymptomaticTB-BrazilPrisons/releases/tag/v1.0.

### Funding Statement

This work was supported by Brazilian Ministry of Health; Brazilian National Council for Scientific and Technological Development [CNPq; 404182/2019-4 to JC] and the U.S. National Institutes of Health [R01 AI130058, R01 AI149620, K24AI182647 to JRA].

