## Supplementary Material for "The role of tuberculosis symptoms in transmission risk to cell contacts in prisons"

**Supplementary Figure 1.** Bivariate analyses of QFT positivity and incarceration factors by prison among all participants (N = 686)

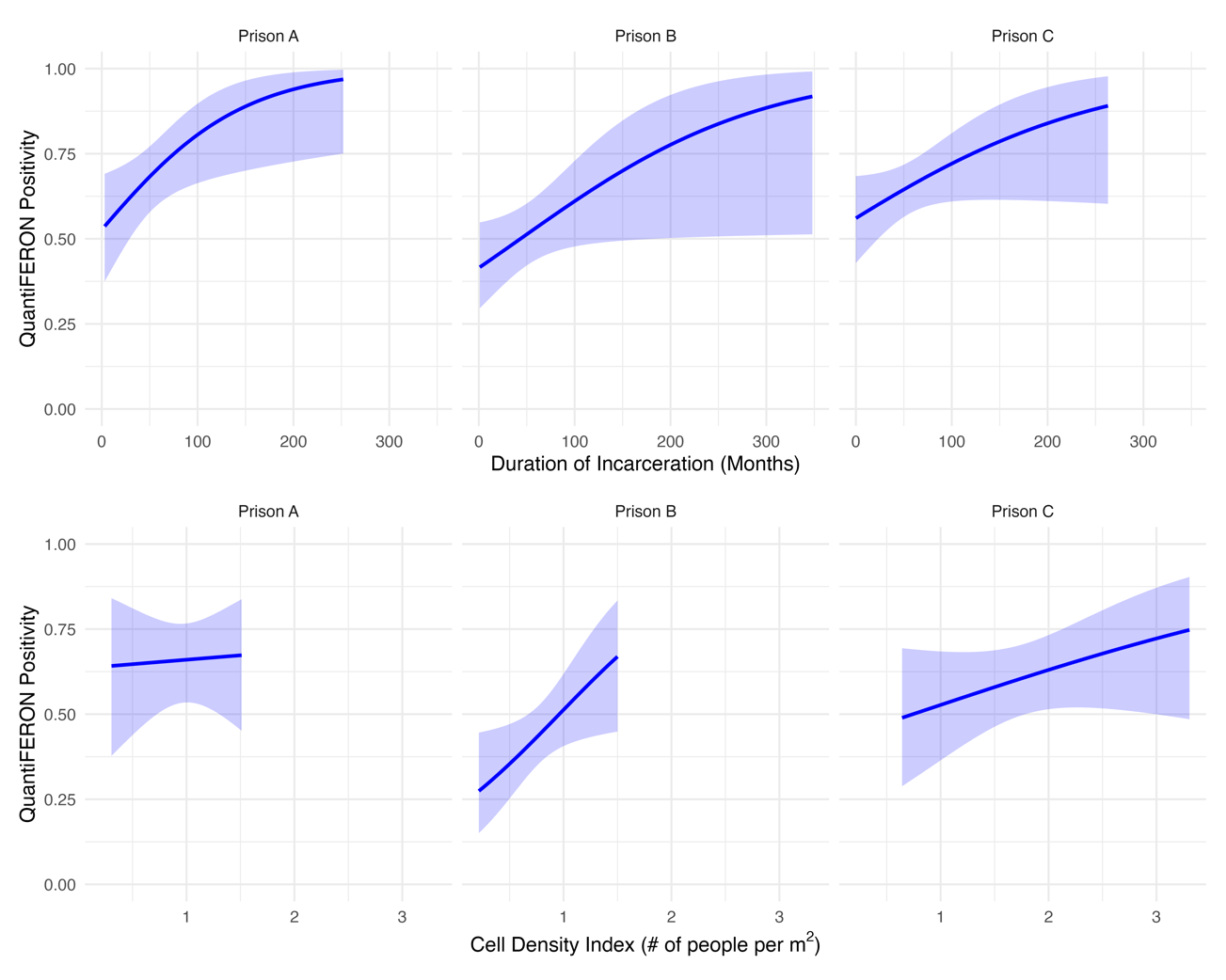

**B**

**A**

**Panel A** - QFT positivity ($\geq$0.35 IU/mL) and incarceration duration by prison, modeled by GAM with logistic regression splines and shaded 95% CI.

**Panel B** - QFT positivity ($\geq$0.35 IU/mL) and cell density index by prison, modeled by GAM with logistic regression splines and shaded 95% CI.

Abbreviations: QFT, QuantiFERON; GAM, Generalized Additive Models; CI, Confidence Intervals.

**Supplementary Table 1.** Sensitivity analyses for QFT positivity evaluating effect of tuberculosis exposure type among individuals with no previous incarceration (N = 686)

| Predictor | **QFT > 0.70 IU/mL** | | | | |  | **QFT > 4.0 IU/mL** | | | | | |
| --- | --- | --- | --- | --- | --- | --- | --- | --- | --- | --- | --- | --- |
|  | aOR | 95% CrI | | | *P* value |  | aOR | 95% CrI | | | *P* value | |
| Type of TB exposure | |  |  |  | | | |  |  | | | |
| Symptomatic vs. Unexposed | 1.97 | 1.22 | 3.19 | | .006 |  | 3.35 | 1.55 | | 7.24 | | .002 |
| Asymptomatic vs. Unexposed | 1.68 | 1.09 | 2.58 | | .019 |  | 3.00 | 1.42 | | 6.36 | | .004 |
| Symptomatic vs. Asymptomatic | 1.17 | 0.69 | 2.00 | | .556 |  | 1.13 | 0.56 | | 2.30 | | .729 |
| Age | 1.00 | 0.98 | 1.01 | | .539 |  | 1.01 | 0.99 | | 1.03 | | .286 |
| Prison |  |  |  | |  |  |  |  | |  | |  |
| A | Reference | | | | |  | Reference | | | | | |
| B | 0.50 | 0.31 | 0.82 | | .006 |  | 0.28 | 0.13 | | 0.61 | | .001 |
| C | 0.67 | 0.34 | 1.30 | | .233 |  | 0.76 | 0.29 | | 1.99 | | .572 |
| Cell density (persons per m^2^) | 1.59 | 0.96 | 2.63 | | .070 |  | 0.87 | 0.43 | | 1.77 | | .696 |
| Months incarcerated | 1.01 | 1.01 | 1.02 | | <.001 |  | 1.01 | 1.01 | | 1.02 | | <.001 |

Abbreviations: QFT, QuantiFERON; aOR, Adjusted Odds Ratio; CrI, Credible Interval.

**Supplementary Table 2.** Sensitivity analyses for QFT positivity evaluating effect of Xpert result among individuals with no previous incarceration (N = 686)

| Predictor | **QFT > 0.70 IU/mL** | | | | | |  | **QFT > 4.0 IU/mL** | | | | | | | | | |
| --- | --- | --- | --- | --- | --- | --- | --- | --- | --- | --- | --- | --- | --- | --- | --- | --- | --- |
|  | aOR | 95% CrI | | | | *P* value | | |  | | aOR | | 95% CrI | | | | *P* value |
| Xpert Result |  |  |  | |  | | | | |  | |  | |  | |  | |
| Very low/Low vs. Negative | 1.54 | 0.94 | | 2.53 | | .088 | | |  | | 2.74 | | 1.26 | | 5.95 | | .011 |
| Medium/High vs. Negative | 2.10 | 1.29 | | 3.42 | | .003 | | |  | | 3.55 | | 1.67 | | 7.54 | | .001 |
| Medium/High vs. Very low/Low | 1.39 | 0.75 | | 2.56 | | .297 | | |  | | 1.31 | | 0.65 | | 2.64 | | .451 |
| Age | 1.00 | 0.98 | | 1.01 | | .582 | | |  | | 1.01 | | 0.99 | | 1.03 | | .256 |
| Prison |  |  | |  | |  | | |  | |  | |  | |  | |  |
| A | Reference | | | | | |  | Reference | | | | | | | | | |
| B | 0.50 | 0.30 | | 0.83 | | .007 | | |  | | 0.29 | | 0.13 | | 0.64 | | .002 |
| C | 0.66 | 0.33 | | 1.30 | | .229 | | |  | | 0.76 | | 0.29 | | 1.95 | | .563 |
| Cell density (persons per m^2^) | 1.65 | 1.00 | | 2.71 | | .050 | | |  | | 0.89 | | 0.42 | | 1.89 | | .757 |
| Months incarcerated | 1.01 | 1.01 | | 1.02 | | <.001 | | |  | | 1.01 | | 1.01 | | 1.02 | | <.001 |

Abbreviations: QFT, QuantiFERON; aOR, Adjusted Odds Ratio; CrI, Credible Interval.
